# Psychosocial mediators for the impact of personal genomic risk information on melanoma prevention and early detection behaviors

**DOI:** 10.64898/2026.05.07.26352695

**Authors:** Sabrina E Wang, David Espinoza, Serigne N Lo, Amelia K Smit, Anne E Cust

## Abstract

**Background:** In the Melanoma Genomics Managing Your Risk Study, access to personal genomic risk testing led to improvements in some melanoma prevention and early detection behaviors.

**Purpose:** We aimed to examine the hypothesized psychosocial mediators of the effects observed in the trial.

**Methods:** Australians of European ancestry without melanoma and aged 18–69 years were recruited via the national Medicare database and randomized to receive personal genomic risk information or usual care (N=1,025). Questionnaires were administered at baseline, 1-month post-intervention, and 12-months post-baseline to assess self-reported prevention and early detection behaviors and psychosocial measures. To identify potential mediators, we first evaluated the intervention’s effect on psychosocial measures and the associations between psychosocial measures and behavioral outcomes. We then estimated the natural indirect effects (NIEs) and their 95% confidence intervals (CIs) to quantify the effects mediated by potential mediators identified.

**Results:** Among participants with high traditional melanoma risk, the intervention’s effect on increased sun protection at 1-month was partially mediated by changes in perceived importance [NIE mean difference (95% CI): 0.02 (0.00, 0.04)] and perceived effectiveness [0.01 (0.00, 0.03)] of sun protection strategies. Among women, the intervention’s effect on increased whole-body skin examinations at 1-month was partially mediated by perceived capability to engage in skin examinations [NIE odds ratio (95% CI): 1.08 (1.00, 1.29)] and perceived control over detecting a future melanoma [1.13 (1.03, 1.32)].

**Conclusions:** The effectiveness of precision prevention and early detection interventions may be enhanced by targeting key psychosocial mediators through tailored communication of personal melanoma risk.

## Introduction

The Melanoma Genomics Managing Your Risk Study is a randomized controlled trial designed to evaluate the impact of providing personal genomic risk information on melanoma prevention and early detection behaviors in a general Australian population. Previous analyses demonstrated that access to personal genomic risk information led to improvements in several behavioral outcomes, most evidently among participants with high traditional melanoma risk scores and among women. The mechanisms underlying these behavioral changes remain unclear.

Understanding the motivations for behavior change is critical, as this knowledge may inform the optimization of prevention and early detection interventions and enhance their public health impact. Health behavior theories offer a framework for explaining how personal genomic risk communication may influence engagement in health behaviors through changes in psychosocial determinants. Specifically, the Protection Motivation Theory [1] and the Health Belief Model[2] propose that perceptions of disease severity and personal susceptibility, along with beliefs regarding the effectiveness and importance of recommended actions, self-efficacy, and perceived barriers, may be key drivers of prevention and early detection behaviors. In a qualitative study, personal melanoma risk communication was observed to help individuals feel better equipped to manage their cancer risk.[3] However, the literature on how personal genomic risk communication leads to behavioral changes remains limited.

In this analysis, we aimed to examine theoretically-derived psychosocial mediators of the effects of personal genomic risk information on melanoma prevention and early detection behaviors observed in the Melanoma Genomics Managing Your Risk Study. We hypothesized that access to personal melanoma risk information would influence some psychosocial determinants, which in turn would mediate the observed behavioral effects of the intervention.

## Methods

### Study design and population

This analysis included participants of the Melanoma Genomics Managing Your Risk Study.[4] Briefly, eligibility for the trial included age 18-69 years at the time of recruitment, no personal history of melanoma, and full or part European ancestry. Recruitment occurred between October 2017 and February 2019, with 1,025 participants completing baseline measures and randomized to receive the intervention (n=513; personalized melanoma risk booklet based on a polygenic risk score, genetic counseling via telephone, educational booklet) or control (n=512; educational booklet). Randomization was stratified by traditional risk phenotype (low, high), sex, state of residence, and age group (18-44, 45-69 years). Hypothesized psychosocial mediators and behavioral outcomes were assessed via questionnaires at baseline, 1-month after intervention (T1), and 12-months after baseline (T2). A total of 994 (97%) participants completed the questionnaire at follow-up 1 and 973 (95%) participants completed the questionnaire at follow-up 2. The detailed study protocol, statistical plan, and main results of the trial are published.[4-6] The methods and results of this present analysis are reported following the Strengthening the Reporting of Observational Studies in Epidemiology Statement for cohort studies.[7]

### Behavioral outcomes

Outcomes of interest were those demonstrating statistically significant between-group differences (p<0.05) in the main trial (Figure 1a). These included: (1) improved sun protection habits at T1 among participants in the high traditional risk group; (2) reduced intentional tanning frequency at T1 among women; (3) increased whole-body skin examination at T1 among women; and (4) reduced sunburn at T2 for all participants.

**Figure 1.**
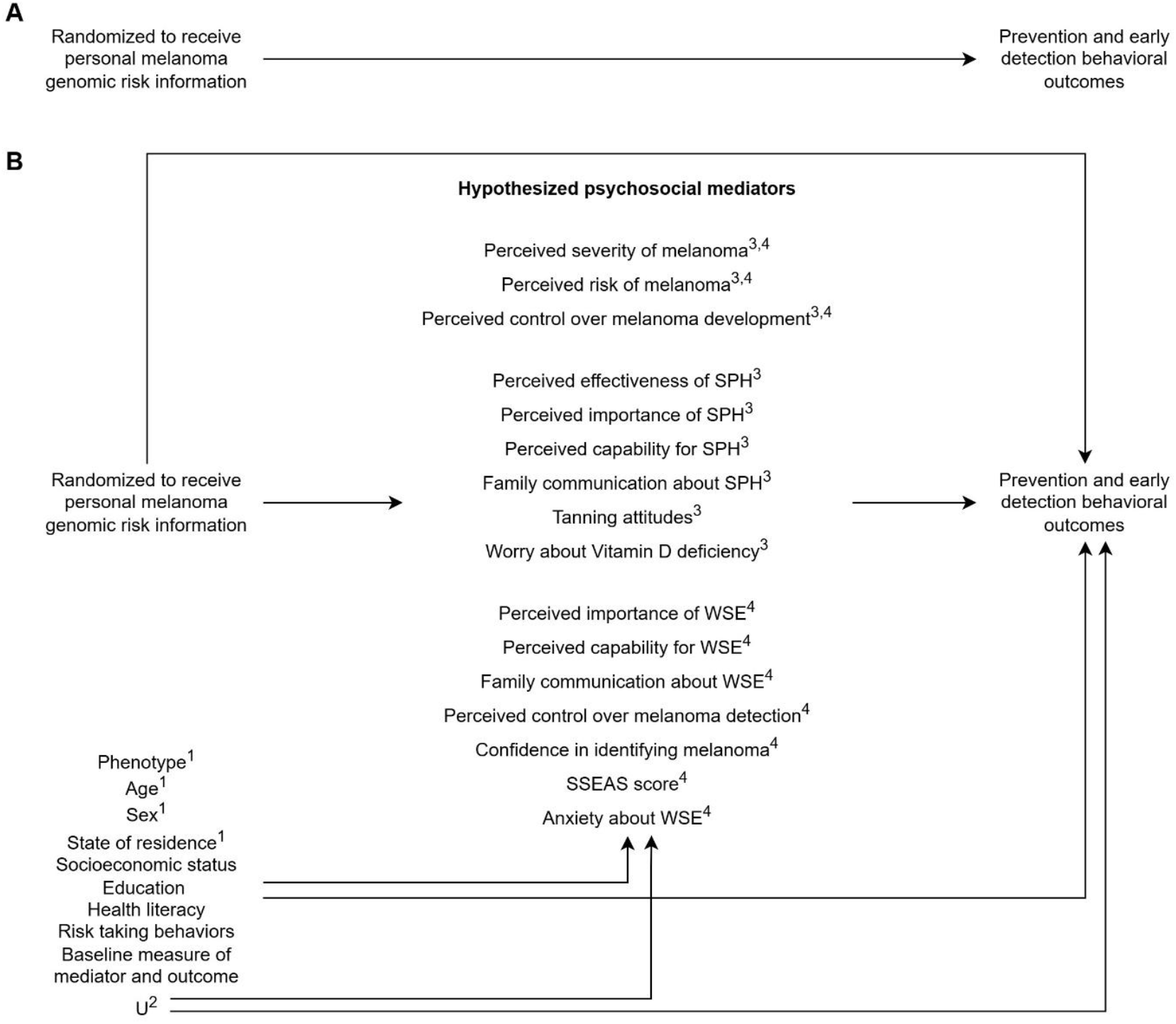
(A) Causal graph illustrating the intervention’s effect on behavioral outcomes in the Melanoma Genomics Managing Your Risk Study trial; (B) Causal graph illustrating the mediation analysis aimed to examine hypothesized psychosocial mediators of the effects observed in the trial

^1^ Stratification factors for randomization

^2^ U denotes unassessed confounding factors

^3^ Hypothesized mediators for the intervention’s effect on sun protection outcomes (i.e., sun protection habits, intentional tanning, and sunburn)

^4^ Hypothesized mediators for the intervention’s effect on early detection behavior (i.e., whole-body skin examination)

SPH = sun protection habits; SSEAS = Skin Self-Examination Attitude Scale; WSE = whole-body skin examination

Sun protection habits index was a validated composite score calculated as the mean of six sun protection habits adopted in the past month (wearing sunscreen, wearing a shirt with sleeves, wearing a hat, staying in the shade, wearing sunglasses, limiting midday sun exposure), each assessed on a 4-point Likert scale (1 = never or rarely, 2 = sometimes, 3 = often, 4 = always).[8] Intentional tanning frequency in the past month was assessed on a 5-point Likert scale (1 = never, 2 = rarely, 3 = sometimes; 4 = often, 5 = always). Whole-body skin examination, self-performed or conducted by a health professional, was dichotomized as ‘yes’ (examination on all or nearly all of the skin) or ‘no’ (all other responses). Sunburn frequency in the past month was collected (0, 1, 2, or 3 or more times) and dichotomized as ‘any’ or ‘none’.

### Hypothesized psychosocial mediators

The development of proposed mediator measures was guided by the constructs of the Protection Motivation Theory[1] and the Health Belief Model[2]. We hypothesized that access to personal melanoma risk information could influence an individual’s perceptions of disease severity, personal susceptibility, as well as their beliefs about the efficacy of, and the ability or barriers to undertake preventive activities, thereby impacting their engagement in health behaviors. The assumed causal structure involving the mediators is illustrated in Figure 1b. For hypothesized mediators referring to sun protection habits, we used the total index score. In an exploratory analysis, we examined individual habits separately (e.g., perceived effectiveness of wearing sunscreen as the hypothesized mediator). For hypothesized mediators referring to whole-body skin examinations, we used the mean Likert scale score of those performed by health professionals and those that were self-performed. In an exploratory analysis, we examined self-performed and health professional-performed skin examinations separately (e.g., perceived capability to engage in skin self-examination as the hypothesized mediator). More details for each mediator measure are published elsewhere.[5] We used psychosocial measures collected 1-month post-intervention (at T1) in our analyses for hypothesized mediators.

### Statistical analysis

We first replicated the intervention-outcome effects of interest (Figure 1a) using linear regression to estimate mean differences and 95% confidence interval (CI) for continuous outcomes (sun protection habits index and tanning frequency) and logistic regression to estimate odds ratios and 95% CI for binary outcomes (whole-body skin examination and sunburn). In the main trial, relative risks were reported for binary outcomes. In contrast, odds ratios were used in the present analysis to address potential convergence issues arising from the multiple adjustment factors planned. Estimates obtained from the main trial and from this analysis are presented side-by-side in Table S1. To identify potential mediators, we then evaluated the intervention-mediator and mediator-outcome associations using linear or logistic regressions, as appropriate (Figure 1b). Analyses were performed using a complete-case approach, given the proportion of missing data for each variable was low (≤5%).

All intervention-outcome, intervention-mediator, and mediator-outcome regression models were adjusted for the following covariates: traditional risk phenotype (low, high), age at baseline, sex (male, female), state of residence (dichotomized based on latitude: SA/VIC/TAS, QLD/NT/WA/NSW/ACT), socioeconomic status (Socio-Economic Indexes for Areas - Index of Relative Socio-Economic Advantage and Disadvantage in quintiles), education (school only, higher education), health literacy (low, high), risk taking behaviors [risk-seeking, risk-neutral, or risk-averse from the ‘Health and Safety’ and ‘Social’ domains from the Domain-Specific Risk-taking scale (DOSPERT)],[9, 10] and baseline measures of mediator or outcome.[5] Regression models assessing mediator-outcome associations additionally included the intervention group as a covariate. Inclusion of these pre-specified covariates ensured appropriate adjustment for potential confounding (for mediator-outcome associations) and consistency with the mediation analysis models.

For psychosocial measures that were associated with both the intervention and a behavioral outcome, we quantified the extent to which the proposed mediator might explain the effect of the intervention on the outcome. Using the PARAMED module in Stata Statistical Software (Release 19),[11] the total effect of the intervention on an outcome was decomposed into natural indirect effect (NIE) and natural direct effect (NDE). Under the counterfactual framework,[12] the NIE represents the mediated effect for an individual and is defined as the difference in the outcome when the individual is set to receive the intervention and their mediator value is set to what it would have been under the intervention, compared with when the individual is set to receive the intervention and their mediator value is set to what it would have been without the intervention. The NDE represents the effect not operating through the mediator and is defined as the difference in outcome when the individual is set to receive the intervention compared with not receiving the intervention, while holding their mediator value at the level it would have been in the absence of the intervention. Bias-corrected bootstraps with 1000 resamples were used to estimate the 95% CIs for the NIE and NDE.

## Results

Baseline characteristics of study participants by study arms are presented for all participants, participants with high traditional melanoma risk, and women separately in Table 1. The intervention-mediator and mediator-outcome associations for each hypothesized psychosocial mediator and behavioral outcome are presented in Table 2. The estimated NIE and NDE for potential mediators identified are presented in Table 3.

**Table 1.** Baseline characteristics of participants by study arm and population subgroup.

|  | All participants (N=1024) |  | High traditional risk (N=505) |  | Women (N=521) |  |
| --- | --- | --- | --- | --- | --- | --- |
|  | Intervention (N=513) | Control (N=511) | Intervention (N=252) | Control (N=253) | Intervention (N=261) | Control (N=260) |
| Age, mean (SD) | 47.1 (14.8) | 46.7 (14.3) | 48.2 (14.0) | 47.9 (13.9) | 46.7 (14.0) | 45.5 (14.5) |
| Sex, N (%) |  |  |  |  |  |  |
| Male | 252 (49.1%) | 251 (49.1%) | 121 (48.0%) | 119 (47.0%) | 0 (0.0%) | 0 (0.0%) |
| Female | 261 (50.9%) | 260 (50.9%) | 131 (52.0%) | 134 (53.0%) | 261 (100.0%) | 260 (100.0%) |
| Phenotype, N (%) |  |  |  |  |  |  |
| High traditional risk | 261 (50.9%) | 258 (50.5%) | 252 (100.0%) | 253 (100.0%) | 130 (49.8%) | 126 (48.5%) |
| Low traditional risk | 252 (49.1%) | 253 (49.5%) | 0 (0.0%) | 0 (0.0%) | 131 (50.2%) | 134 (51.5%) |
| State of residence, N (%) |  |  |  |  |  |  |
| SA/VIC/TAS | 197 (38.4%) | 197 (38.6%) | 82 (32.5%) | 80 (31.6%) | 101 (38.7%) | 111 (42.7%) |
| QLD/NT/WA/NSW/ACT | 316 (61.6%) | 314 (61.4%) | 170 (67.5%) | 173 (68.4%) | 160 (61.3%) | 149 (57.3%) |
| Socioeconomic status, N (%) |  |  |  |  |  |  |
| 1 (most disadvantaged) | 98 (19.3%) | 107 (21.2%) | 41 (16.4%) | 50 (20.1%) | 47 (18.1%) | 53 (20.7%) |
| 2 | 115 (22.6%) | 89 (17.7%) | 59 (23.6%) | 44 (17.7%) | 65 (25.1%) | 42 (16.4%) |
| 3 | 97 (19.1%) | 102 (20.2%) | 54 (21.6%) | 56 (22.5%) | 48 (18.5%) | 56 (21.9%) |
| 4 | 109 (21.4%) | 95 (18.8%) | 49 (19.6%) | 47 (18.9%) | 54 (20.8%) | 51 (19.9%) |
| 5 | 90 (17.7%) | 111 (22.0%) | 47 (18.8%) | 52 (20.9%) | 45 (17.4%) | 54 (21.1%) |
| Education, N (%) |  |  |  |  |  |  |
| School | 120 (23.4%) | 119 (23.3%) | 52 (20.6%) | 58 (22.9%) | 47 (18.0%) | 57 (21.9%) |
| Higher education | 393 (76.6%) | 392 (76.7%) | 200 (79.4%) | 195 (77.1%) | 214 (82.0%) | 203 (78.1%) |
| Health literacy, N (%) |  |  |  |  |  |  |
| Low | 66 (12.9%) | 49 (9.6%) | 30 (11.9%) | 21 (8.3%) | 20 (7.7%) | 17 (6.5%) |
| High | 447 (87.1%) | 462 (90.4%) | 222 (88.1%) | 232 (91.7%) | 241 (92.3%) | 243 (93.5%) |
| Health risk-taking, N (%) |  |  |  |  |  |  |
| Risk-averse | 64 (12.5%) | 71 (13.9%) | 28 (11.1%) | 35 (13.8%) | 48 (18.4%) | 48 (18.5%) |
| Risk-neutral | 374 (72.9%) | 351 (68.7%) | 193 (76.6%) | 181 (71.5%) | 189 (72.4%) | 187 (71.9%) |
| Risk-seeking | 75 (14.6%) | 89 (17.4%) | 31 (12.3%) | 37 (14.6%) | 24 (9.2%) | 25 (9.6%) |
| Social risk-taking, N (%) |  |  |  |  |  |  |
| Risk-averse | 96 (18.7%) | 91 (17.8%) | 33 (13.1%) | 54 (21.3%) | 57 (21.8%) | 43 (16.5%) |
| Risk-neutral | 315 (61.4%) | 327 (64.0%) | 169 (67.1%) | 156 (61.7%) | 153 (58.6%) | 164 (63.1%) |
| Risk-seeking | 102 (19.9%) | 93 (18.2%) | 50 (19.8%) | 43 (17.0%) | 51 (19.5%) | 53 (20.4%) |

**Table 2.** Intervention-mediator and mediator-outcome associations for hypothesized psychosocial mediators by outcome of interest.

|  | Intervention-mediator<sup>1</sup> | Mediator-outcome<sup>1</sup> |
| --- | --- | --- |
| **SPH at T1 among the high traditional risk group (N=482)** | Mean difference (95% CI) | Mean difference (95% CI) |
| Perceived severity of melanoma | -0.02 (-0.13, 0.09) | 0.02 (-0.04, 0.09) |
| Perceived absolute risk of melanoma | **-13.85 (-17.47, -10.23)** | **-0.00 (-0.00, -0.00)** |
| Perceived relative risk of melanoma | -0.10 (-0.23, 0.02) | -0.01 (-0.06, 0.04) |
| Perceived control over melanoma development | -0.04 (-0.19, 0.11) | 0.04 (-0.01, 0.08) |
| Perceived effectiveness of SPH | **0.13 (0.03, 0.23)** | **0.10 (0.03, 0.16)** |
| Perceived importance of SPH | **0.09 (0.02, 0.17)** | **0.20 (0.11, 0.29)** |
| Perceived capability for SPH | 0.02 (-0.06, 0.10) | **0.15 (0.07, 0.23)** |
| Family communication on SPH | 0.06 (-0.10, 0.22) | **0.16 (0.12, 0.20)** |
| Tanning attitudes (Pro-tan score) | -0.22 (-0.83, 0.40) | **-0.01 (-0.02, -0.00)** |
| Worry about Vitamin D deficiency | -0.02 (-0.10, 0.05) | -0.06 (-0.15, 0.03) |
| **Intentional tanning at T1 among women (N=499)** | Mean difference (95% CI) | Mean difference (95% CI) |
| Perceived severity of melanoma | -0.03 (-0.14, 0.08) | **0.08 (0.00, 0.16)** |
| Perceived absolute risk of melanoma | **-14.46 (-17.88, -11.04)** | 0.00 (-0.00, 0.00) |
| Perceived relative risk of melanoma | **-0.17 (-0.29, -0.04)** | 0.00 (-0.07, 0.07) |
| Perceived control over melanoma development | 0.09 (-0.04, 0.23) | 0.01 (-0.06, 0.07) |
| Perceived effectiveness of SPH | **0.13 (0.03, 0.22)** | -0.03 (-0.12, 0.06) |
| Perceived importance of SPH | 0.06 (-0.00, 0.13) | **-0.15 (-0.27, -0.02)** |
| Perceived capability for SPH | 0.01 (-0.07, 0.09) | -0.09 (-0.20, 0.02) |
| Family communication on SPH | 0.12 (-0.06, 0.29) | 0.04 (-0.01, 0.09) |
| Tanning attitudes (Pro-tan score) | -0.42 (-1.09, 0.25) | **0.03 (0.01, 0.04)** |
| Worry about Vitamin D deficiency | -0.06 (-0.14, 0.01) | 0.03 (-0.08, 0.15) |
| **WSE at T1 among women (N=499)** | Mean difference (95% CI) | Odds ratios (95% CI) |
| Perceived severity of melanoma | -0.03 (-0.14, 0.08) | 0.94 (0.66, 1.34) |
| Perceived absolute risk of melanoma | **-14.46 (-17.88, -11.04)** | 1.00 (0.99, 1.01) |
| Perceived relative risk of melanoma | **-0.17 (-0.29, -0.04)** | 0.90 (0.66, 1.23) |
| Perceived control over melanoma development | 0.09 (-0.04, 0.23) | 1.11 (0.82, 1.52) |
| Perceived importance of WSE | 0.06 (-0.02, 0.15) | 1.60 (0.95, 2.71) |
| Perceived capability for WSE | **0.10 (0.01, 0.19)** | **2.33 (1.39, 3.92)** |
| Family communication on WSE | 0.03 (-0.14, 0.21) | **1.87 (1.48, 2.37)** |
| Perceived control over melanoma detection | **0.21 (0.07, 0.35)** | **1.91 (1.39, 2.63)** |
| Confidence identifying melanoma | 0.08 (-0.07, 0.22) | **2.24 (1.65, 3.05)** |
| SSEAS score | 0.35 (-0.31, 1.01) | **1.23 (1.13, 1.33)** |
| Anxiety about WSE | -0.14 (-0.28, 0.01) | 0.91 (0.68, 1.21) |
| **Sunburn at T2 (N=961)** | Mean difference (95% CI) | Odds ratios (95% CI) |
| Perceived severity of melanoma | 0.01 (-0.07, 0.08) | 1.00 (0.70, 1.42) |
| Perceived absolute risk of melanoma | **-13.89 (-16.37, -11.41)** | **1.01 (1.00, 1.02)** |
| Perceived relative risk of melanoma | -0.08 (-0.17, 0.02) | 1.28 (0.96, 1.69) |
| Perceived control over melanoma development | 0.01 (-0.09, 0.11) | 0.84 (0.65, 1.09) |
| Perceived effectiveness of SPH | 0.06 (-0.01, 0.13) | 0.83 (0.58, 1.18) |
| Perceived importance of SPH | 0.03 (-0.02, 0.08) | 0.89 (0.54, 1.45) |
| Perceived capability for SPH | -0.00 (-0.06, 0.05) | **0.63 (0.40, 0.99)** |
| Family communication on SPH | 0.08 (-0.04, 0.20) | 1.01 (0.81, 1.26) |
| Tanning attitudes (Pro-tan score) | -0.09 (-0.53, 0.35) | 1.05 (0.99, 1.11) |
| Worry about Vitamin D deficiency | -0.03 (-0.08, 0.02) | 1.04 (0.63, 1.72) |

<sup>1</sup>Estimates in bold font indicates p<0.05
CI = confidence interval; SPH = sun protection habits; SSEAS = Skin Self-Examination Attitude Scale; WSE = whole-body skin examination

**Table 3.** Estimated natural indirect effects for potential mediators of intervention’s effect on melanoma prevention or early detection behavioral outcomes.

| Outcome | Mediator |  |  |  |  |
| --- | --- | --- | --- | --- | --- |
| **SPH at T1 among the high traditional risk group (N=482)** |  | **TE<sup>1</sup>** | **0.13 (0.06, 0.20)** |  |  |
|  | Perceived importance of SPH | NIE<sup>1</sup> | 0.02 (0.00, 0.04)<sup>3</sup> | NDE<sup>1</sup> | 0.11 (0.03, 0.18)<sup>3</sup> |
|  | Perceived effectiveness of SPH | NIE<sup>1</sup> | 0.01 (0.00, 0.03)<sup>3</sup> | NDE<sup>1</sup> | 0.12 (0.05, 0.19)<sup>3</sup> |
| **WSE at T1 among women (N=499)** |  | **TE<sup>2</sup>** | **1.83 (1.15, 2.91)** |  |  |
|  | Perceived capability for WSE | NIE<sup>2</sup> | 1.08 (1.00, 1.29)<sup>3</sup> | NDE<sup>2</sup> | 1.67 (0.94, 2.64)<sup>3</sup> |
|  | Perceived control over melanoma detection | NIE<sup>2</sup> | 1.13 (1.03, 1.32)<sup>3</sup> | NDE<sup>2</sup> | 1.55 (0.95, 2.52)<sup>3</sup> |
| **Sunburn at T2 (N=961)** |  | **TE<sup>2</sup>** | **0.62 (0.41, 0.94)** |  |  |
|  | Perceived absolute risk of melanoma | NIE<sup>2</sup> | 0.92 (0.75, 1.11)<sup>3</sup> | NDE<sup>2</sup> | 0.66 (0.42, 1.05)<sup>3</sup> |

<sup>1</sup> Estimated mean difference and 95% confidence interval
<sup>2</sup> Estimated odds ratios and 95% confidence interval
<sup>3</sup> Bias-corrected bootstrapped 95% confidence intervals with 1000 replications
NDE = natural direct effect; NIE = natural indirect effect; SPH = sun protection habits; TE = total effect; WSE = whole-body skin examination

### Sun protection habits at T1 among participants with high traditional melanoma risk

Among participants with high traditional melanoma risk, access to personal melanoma genomic risk information improved sun protection habits at T1 [mean difference in total index score 0.13; 95% CI 0.06, 0.20]. The intervention increased perceived effectiveness (mean difference in average combined score 0.13; 95% CI 0.03, 0.23) and perceived importance (0.09; 95% CI 0.02, 0.17) of sun protection habits in reducing melanoma risk at T1. Perceived effectiveness (mean difference in average combined score 0.10; 95% CI 0.03, 0.16) and perceived importance (0.20; 95% CI 0.11, 0.29) were positively associated with sun protection habits at T1. In mediation analysis, perceived importance (NIE mean difference 0.02; 95% CI 0.00, 0.04) and perceived effectiveness (0.01; 0.00, 0.03) of sun protection habits partially mediated the intervention’s effect on increased sun protection behaviors.

### Intentional tanning at T1 among women

Among women, access to personal melanoma genomic risk information reduced intentional tanning at T1 (mean difference -0.12; 95% CI -0.22, -0.02). No psychosocial factor was associated with both the intervention and intentional tanning frequency, thus no further mediation analysis was performed.

### Whole-body skin examination at T1 among women

Among women, access to personal melanoma genomic risk information increased whole-body skin examinations at T1 [odds ratio (OR) 1.83; 95% CI 1.15, 2.91]. The intervention increased perceived capability to engage in whole-body skin examinations (mean difference 0.13; 95% CI 0.02, 0.24) and perceived control over detecting a future melanoma early in its development (0.21; 95% CI 0.07, 0.35) at T1. Perceived capability to engage in whole-body skin examinations (OR 2.33; 95% CI 1.39, 3.92) and perceived control over detecting a future melanoma (1.91; 1.39, 2.63) were positively associated with whole-body skin examinations at T1. In mediation analysis, perceived capability to engage in skin examinations (NIE OR 1.08; 95% CI 1.00, 1.29) and perceived control over detecting a future melanoma (1.13; 1.03, 1.32) partially mediated the intervention’s effect on increased whole-body skin examinations.

### Sunburn at T2

Access to personal melanoma genomic risk information reduced self-reported sunburn incidence at T2 (OR 0.62; 95% CI 0.41, 0.94), most evidently among participants in the high traditional risk group (OR 0.41; 95% CI 0.22, 0.76) and among women (OR 0.36; 95% CI 0.18, 0.70). The intervention reduced perceived absolute risk of melanoma (mean difference on a 0 to 100 scale - 13.89; 95% CI -16.37, -11.41), which was positively associated with odds of sunburn (OR 1.01; 95% CI 1.00, 1.02). In mediation analysis, there was no evidence that perceived absolute risk of melanoma mediated the intervention’s effect on reduced sunburn (NIE OR 0.92; 95% CI 0.75, 1.11). Consistent results were observed among participants in the high traditional risk group (perceived absolute risk of melanoma NIE OR 1.05; 95% CI 0.74, 1.55) and among women (0.94; 0.76, 1.02).

### Exploratory analyses

In the exploratory analysis evaluating individual sun protection habits and types of skin examination separately, we identified several potential mediators: (1) the intervention increased perceived effectiveness of protective clothing (mean difference 0.17; 95% CI 0.04, 0.30), which was positively associated with sun protection habits (0.08; 0.03, 0.13) at T1 among participants in the high traditional risk group, the NIE was 0.01 (95% CI 0.00, 0.03) indicating partial mediation; (2) the intervention increased perceived importance of wearing long pants (mean difference 0.13; 95% CI 0.01, 0.26), which was positively associated with sun protection habits (0.09; 0.04, 0.15) at T1 among participants in the high traditional risk group, the NIE was 0.01 (95% CI 0.00, 0.04) indicating partial mediation; (3) the intervention increased perceived effectiveness of limiting midday sun exposure (mean difference 0.11; 95% CI 0.00, 0.23), which was inversely associated with intentional tanning (-0.08; -0.16, -0.00) at T1 among women, the NIE was 0.00 (95% CI -0.02, 0.01) indicating no mediation; (4) the intervention increased perceived capability to engage in skin self-examinations (mean difference 0.13; 95% CI 0.02, 0.24), which was positively associated with whole-body skin examinations (OR 1.97; 95% CI 1.31, 2.95) at T1 among women, the NIE OR was 1.12 (95% CI 1.01, 3.17) indicating partial mediation; (5) the intervention increased perceived importance of staying in the shade (mean difference 0.10; 95% CI 0.00, 0.19), which was inversely associated with sunburn (OR 0.52; 95% CI 0.29, 0.95) at T2 among women, the NIE OR was 0.94 (95% CI 0.76, 1.02) indicating no mediation.

## Discussion

We evaluated the hypothesized psychosocial mediators for four behavioral outcomes observed in the Melanoma Genomics Managing Your Risk Study. Our key findings were: (1) increased perceived importance and increased perceived effectiveness of sun protection habits partly mediated the effect of access to personal melanoma risk information on increased sun protection behaviors among participants with high traditional melanoma risk; (2) increased perceived capability to engage in skin examinations and increased perceived control over detecting a future melanoma partly mediated the intervention’s effect on increased whole-body skin examinations among women; (3) we did not find any psychosocial mediators for the intervention’s effect on reduced intentional tanning among women; (4) perceived absolute risk of melanoma was identified as a potential mediator for the intervention’s effect on reduced self-reported sunburn for all study participants, but results from the mediation analysis suggested it did not mediate the observed effect.

The mediation analysis assumes there are no unmeasured confounding of intervention-outcome, intervention-mediator, and mediator-outcome relationships, and no intervention-induced mediator-outcome confounding.[12] Given the intervention was randomized, the assumptions of unconfounded intervention-outcome and intervention-mediator relationships were satisfied. For mediator-outcome associations, our analysis accounted for a comprehensive set of baseline demographic, social, and behavioral factors, although residual confounding from unmeasured factors cannot be ruled out.

To ensure the intervention preceded psychosocial and behavioral changes, we used prospective data for the analyses of intervention-mediator and intervention-outcome associations. For the analysis of association between psychosocial measures and behavioral outcomes observed at T1, cross-sectional data were used. We assumed psychosocial changes preceded behavioral changes, acknowledging that reverse causation remains possible. For the analysis of association between psychosocial measures and sunburn observed at T2, prospective data were used.

It is difficult to disentangle the independent effects of access to personalized genomic risk information and genetic counseling on psychosocial and behavioral changes. Alongside delivery of a personalized booklet with their risk information and how they could manage their risk, participants in the intervention arm received one-on-one telephone-based genetic counseling (average duration 7 mins, range 2-25 mins), during which they were offered the opportunity to ask questions regarding sun protection and skin examination after receiving their genomic risk results. The most prevalent discussion topics during the counselling call related to past sun exposure and past and current sun habits,[13] and these conversations may also have affected participants’ psychosocial measures. Qualitative interviews with a subgroup of participants from the main trial indicated that the effect contributed by telephone counseling may have varied by genomic risk level. Some participants who received average or low genomic risk results perceived the telephone counseling as providing limited additional benefit beyond the personalized booklet.[13]

As observed in the main trial, the impact of providing personal genomic risk information on health-related behavioral outcomes varied across population subgroups.[4] Although reductions in sunburn were observed among all participants, other prevention and early detection behaviors were evident only within specific subgroups, thereby limiting the generalizability of the findings to the whole population. Notably, these subgroups, namely participants with high traditional risk and women, who were more likely than men to engage in intentional tanning,[4] represent populations at higher risk for melanoma, who may benefit the most from access to personal genomic risk information. Personal risk communication strategies targeting these populations may thus be further optimized by addressing the psychosocial mediators identified in this study.

Our findings indicate that perceived importance and perceived effectiveness of sun protection habits, most evidently for wearing protective clothing, may partially mediate the effect of access to personal melanoma risk testing on increased sun protection behaviors among participants with high traditional melanoma risk. The estimated NIE was small, suggesting there are additional unmeasured factors mediating the observed effect, such as participation in the trial itself and engagement with a genetic counselor. Our findings are in line with prior research among individuals with a strong family history of melanoma, where genetic testing for *CDKN2A/p16* was associated with sustained improvements in sun protection behaviors from one month up to two years post-testing, with the greatest improvements observed for wearing photoprotective clothing.[14, 15] Similarly, a previous randomized trial on providing personal melanoma genomic risk information to individuals with a strong family history reported a statistically significant difference between the intervention and control arm in wearing long-sleeved shirts.[16] Collectively, the existing body of evidence suggests that compared to other sun protection behaviors, such as regular sunscreen reapplication or consistent shade seeking, wearing protective clothing may represent a comparatively easier sun protection behavior to adopt.

Access to personal melanoma risk information increased perceived capability to engage in skin examinations and perceived control over detecting a future melanoma, which partially mediated the intervention’s effect on increased whole-body skin examinations among women. These findings are consistent with existing evidence suggesting that increased knowledge and confidence may facilitate skin examination behaviors. In a qualitative study, 19 of 30 participants reported satisfaction with genomic testing, with gaining new knowledge about their melanoma risk or about preventive behaviors identified as a key reason.[3] Participants across all genomic risk groups who perceived knowledge gain also reported feeling better equipped to manage their skin cancer risk.[3] Similarly, a U.S. population-based cross-sectional survey found that higher levels of melanoma knowledge and confidence in performing skin self-examination were positively associated with skin self-examination behavior.[17]

Receiving personal genomic melanoma risk information led to reduced perceived absolute risk of melanoma, in line with existing evidence that most individuals overestimate their risk of cancer.[18] Reduced perceived absolute risk of melanoma was also identified as a potential mediator of the intervention’s effect on reduced self-reported sunburn, but results from the mediation analysis suggest perceived risk of melanoma did not mediate the observed effect. Similarly, there was no evidence that the intervention’s effect on reduced intentional tanning among women was mediated by any of the psychosocial mediators evaluated, suggesting the effect of the intervention on observed outcomes was mediated by unmeasured pathways. Given sunburn and intentional tanning are key risk factors for melanoma, future studies aimed to evaluate other possible mediating pathways for intervention’s effect on reduced intentional tanning and sunburn are warranted.

## Data Availability

The Study Protocol (DOI: 10.1016/j.cct.2018.05.014) and Statistical Analysis Plan (DOI: 10.1186/s13063-020-04351-w) for the main trial have been published. De-identified data from this study are not available in a public archive. Requests for obtaining de-identified data should be addressed to the corresponding author. Analytic code used to conduct the analyses presented in this study are not available in a public archive. They may be made available by emailing the corresponding author. Materials used to conduct the study are not publicly available.

## Acknowledgements

We acknowledge other Investigators of the Melanoma Genomics Managing Your Risk Study: Ainsley J. Newson, Rachael L. Morton, Michael Kimlin, Louise Keogh, Matthew H. Law, Judy Kirk, Suzanne J. Dobbinson, Peter A. Kanetsky, Graham J. Mann, Hugh Dawkins, Jacqueline Savard, Kate Dunlop, Lyndal Trevena, Mark Jenkins, Martin Allen, Phyllis Butow, Sarah Wordsworth, Cynthia Low.

